# Investigating neurophysiological effects of a short course of tDCS for cognition in schizophrenia: a target engagement study

**DOI:** 10.1101/2022.03.02.22271807

**Authors:** Kate E Hoy, Hannah Coyle, Kirsten Gainsford, Aron T Hill, Neil W Bailey, Paul B Fitzgerald

## Abstract

**Background:** Cognitive impairment is highly prevalent in schizophrenia and treatment options are severely limited. Development of effective treatments will rely on successful engagement of biological targets. There is growing evidence that the cognitive impairments in schizophrenia are related to impairments in prefrontal cortical inhibition and dysfunctional cortical oscillations.

**Methods:** In the current study we sought to investigate whether a short course of transcranial Direct Current Stimulation (tDCS) could modulate these pathophysiological targets. Thirty participants with schizophrenia were recruited and underwent neurobiological assessment (Transcranial Magnetic Stimulation combined with EEG [TMS-EEG] and task-related EEG) and assessment of cognitive functioning (n-back task and the MATRICS Consensus Cognitive Battery). Participants were then randomized to receive 5 sessions of either active or sham anodal tDCS to the left prefrontal cortex. Twenty-four hours after the last tDCS session participants repeated the neurobiological and cognitive assessments. Neurobiological outcome measures were TMS-evoked potentials (TEPs), TMS-related oscillations and oscillatory power during a 2-back task. Cognitive outcome measures were *d* prime and accurate reaction time on the 2-back and MATRICS scores.

**Results:** Following active tDCS there was a significant reduction in the N40 TEP amplitude in the left parietal occipital region. There were no other significant changes.

**Conclusions:** Future interrogation of evidence based therapeutic targets in large scale RCTs is required.

## 1. Introduction

Cognitive impairment is a core feature of schizophrenia, with attention/working memory, processing speed, verbal memory and executive functioning the main domains impacted (Bora et al., 2010; Insel, 2010). Cognitive symptoms are associated with significant functional disability, yet to date therapeutic approaches have been insufficient (Abbot, 2010; Green et al., 2004; Shamsi et al., 2011). Development of effective treatments for the cognitive impairment in schizophrenia will depend upon being able to effectively target pathophysiology related to these symptoms (Insel, 2010). Past research has repeatedly implicated the connected pathophysiologies of impaired γ-aminobutyric acid (GABA)-ergic function (Frankle et al., 2015), reduced prefrontal cortical inhibition (Lisman, 2012; Sohal and Rubenstein., 2019) and dysregulated oscillatory activity in the cognitive symptoms of schizophrenia (Gonzalez-Burgos and Lewis, 2008; Schmiedt et al., 2005). These pathophysiologies, in particular cortical inhibition and oscillatory activity, have been shown to be amenable to modulation by non-invasive brain stimulation (Cirillo et al., 2017; Hoy et al., 2015; Veniero et al., 2015).

Non-invasive brain stimulation uses electricity to modulate excitation and inhibition, as well as neuronal firing patterns (Cirillo et al., 2017). This is achieved in a number of ways, including via direct electrical stimulation through electrodes placed on the scalp (transcranial Electrical Stimulation [tES]), or by inducing an electrical current in the brain via the application of highly focussed magnetic fields to the head (Transcranial Magnetic Stimulation [TMS]) (Hoy and Fitzgerald, 2010). A number of studies have investigated non-invasive brain stimulation for improving cognition in schizophrenia, with mixed findings (Hasan et al., 2016; Jiang et al., 2019; Kostova et al., 2020; Mervis et al, 2017; Narita et al., 2020). These trials have used largely diverse approaches and parameters, often lacking neurobiological mechanistic hypotheses; particularly those using tES (Kostova et al., 2020; Narita et al., 2020). Addressing these limitations is crucial for advancing treatment development.

We have conducted, to the best of our knowledge, the only preliminary work looking at optimal tES parameters, and effects on posited therapeutic targets, with respect to the cognitive symptoms of schizophrenia (Hoy et al., 2014, 2015, 2016). We initially found that 2mA anodal transcranial Direct Current Stimulation (tDCS), as compared to 1mA and sham, was effective in modulating cortical oscillations and improving working memory; positing that tDCS may have impacted GABA-ergic activity thus producing the required cortical environment for optimal oscillatory activity (Hoy et al., 2014, 2015). Indeed, anodal tDCS has been shown to impact GABA-ergic activity and has been hypothesised to restore the cortical excitation and inhibition balance thought essential for generation and modulation of oscillations (Krause et al., 2013).

We subsequently investigated whether transcranial Alternating Current Stimulation (tACS) was more effective than tDCS or sham in improving working memory in schizophrenia (Hoy et al., 2016); finding that despite its proposed mechanism of modulating endogenous oscillations tACS was not more effective. Collectively these findings suggest that 2mA anodal tDCS may be the most promising approach for cognitive impairment in schizophrenia. This is consistent with past research indicating that the abnormalities in oscillatory activity in schizophrenia are likely to be a downstream consequence of impaired prefrontal cortical inhibition (Gonzalez-Burgos and Lewis, 2008;). Therefore, it is likely that an alteration of impaired GABA-ergic activity, potentially achievable via tDCS, could help restore oscillatory activity in schizophrenia with subsequent improvement of cognition (Hoy et al., 2016).

In the current study we sought to directly investigate whether repeated sessions of anodal tDCS in participants with schizophrenia were able to engage proposed therapeutic targets and subsequently improve cognition. In order to identify the specific neurophysiological targets of intervention we conducted an analysis of the baseline data from the current trial, which included the 30 participants with schizophrenia described here as well as 27 healthy controls (Hoy et al., 2021). We identified reduced prefrontal cortical reactivity (a posited measure of cortical inhibition and excitation) and reduced theta oscillatory activity (both in response to TMS and task-related) as possible markers of impaired cognition in schizophrenia (Hoy et al., 2021). Cortical reactivity was assessed using combined TMS-EEG and indexed via TMS-Evoked Potentials (TEPs) (Hoy et al, 2021). Therefore, for the current trial, we hypothesised that active stimulation, compared to sham, would lead to increased prefrontal cortical reactivity, enhanced theta oscillatory activity (both TMS-related and task-related) and improved cognitive performance.

## 2. Methods

### 2.1 Participants

A total of 30 participants with schizophrenia were recruited to the study, see Table One for demographic and clinical characteristics. Four participants withdrew prior to completing the full simulation course, leaving 26 participants who completed both the pre- and post-stimulation assessments (active = 13, sham = 13). Final numbers for analysis across all variables, and reasons for missing data, are provided in Figure One.

**Figure 1:**
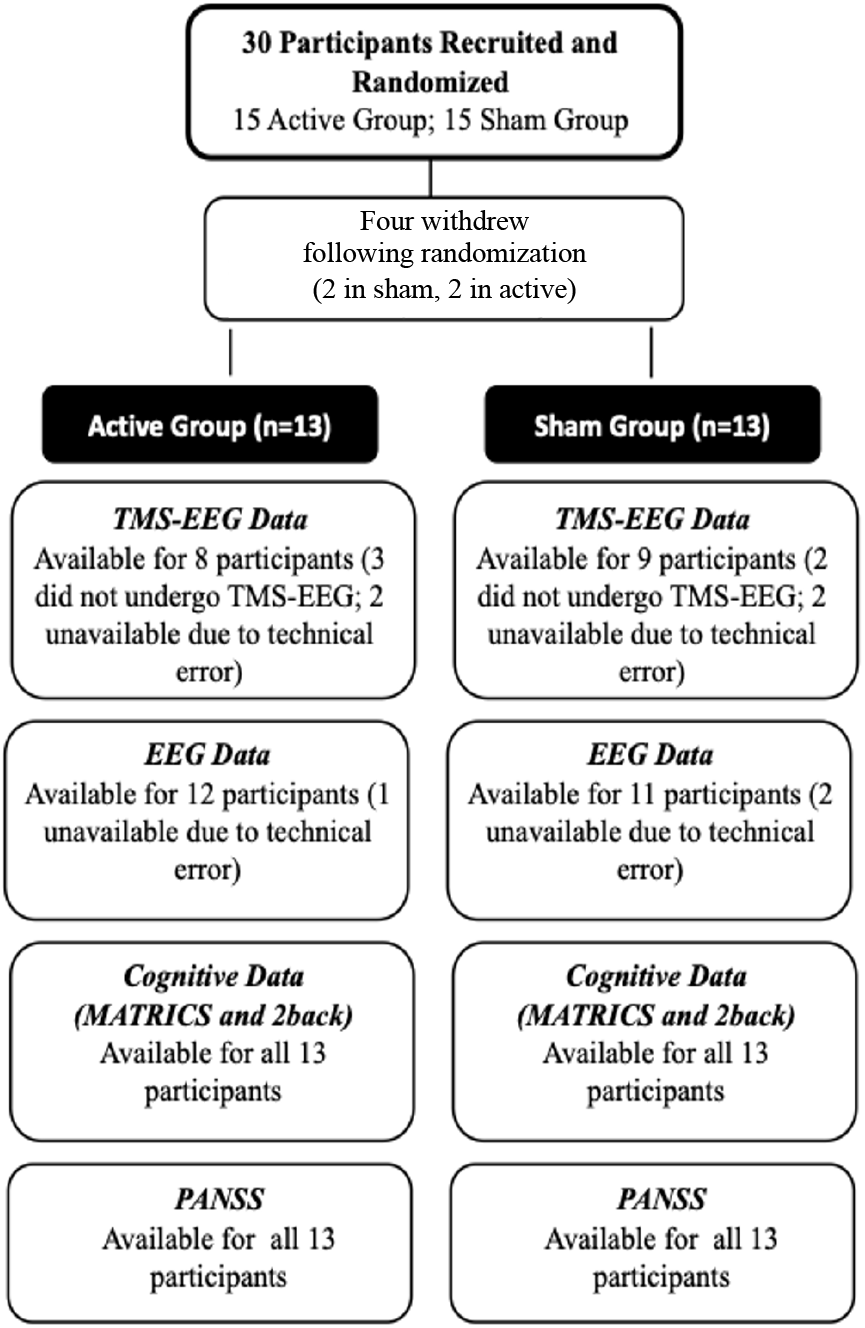
Participant algorithm showing final numbers for analysis across all variables, and reasons for missing data.

**Table 1.** Demographic and Clinical Characteristics for the active and sham tDCS groups

|  | **Active tDCS Group**<br>**(n = 15)** | **Sham tDCS Group**<br>**(n=15)** |  |  |
| --- | --- | --- | --- | --- |
| ***Demographics*** | **Mean ± sd** | **Mean ± sd** | **Test statistic** | **p-value** |
| Age | 45.60 ± 2.34 | 46.20 ± 6.11 | t = 0.21 | p = 0.83 |
| Gender (f/m) | (4/11) | (6/9) | $\chi^2 = 0.60$ | p = 0.44 |
| Handedness (l/r/both) | (13/1/1) | (13/1/1) | $\chi^2 = 0.00$ | p = 1.00 |
| ***Clinical Data*** |  |  |  |  |
| Years since diagnosis | 18.46 ± 8.80 | 21.00 ± 9.39 | t = 0.74 | p = 0.46 |
| PANSS positive | 18.87 ± 6.48 | 18.67 ± 3.88 | t = 0.10 | p = 0.91 |
| PANSS negative | 17.27 ± 3.59 | 17.33 ± 6.95 | t = 0.03 | p = 0.97 |
| PANSS general | 38.60 ± 7.57 | 36.60 ± 8.88 | t = 0.66 | p = 0.51 |
| PANSS total | 74.73 ± 14.16 | 72.60 ± 14.52 | t = 0.40 | p = 0.68 |
| CPZ equivalent (mg/day) | 718.33 ± 519.48 | 755.00 ± 915.89 | t = 0.13 | p = 0.89 |

Diagnosis was confirmed using the Mini-International Neuropsychiatric Interview (MINI; Sheehan et al., 1998). The MINI was conducted by trained research staff experienced in its use (KH/KG/HC). Twenty-nine of the 30 schizophrenia participants were regularly taking antipsychotic medications. Eighteen of the 29 were taking monotherapy (i.e.,4 olanzapine, 3 fluphenazine, 2 aripiprazole, 2 clozapine, 2 amisulpride, 1 risperidone, 1 ziprasidone, 1 paliperidone, 1 trifluoperazine, 1 haliperidol); and 11 were on polytherapy (i.e., 1 olanzapine + clozapine, 1 aripiprazole + quetiapine, 1 risperidone + quetiapine, 1 aripiprazole + zuclopenthixol, 1 clozapine + zuclopenthixol, 1 aripiprazole + clozapine, 1 quetiapine + lurasidone, 1 clozapine + haloperidol, 1 olanzapine + aripiprazole; 1 olanzapine + quetiapine + clotiapine, 1 clozapine + aripiprazole). Eleven participants were additionally taking antidepressant medication (i.e. 7 SSRIs, 2 SNRIs, 2 NaSSA); 10 were also taking mood stabilizers (i.e. 6 lithium, 4 sodium valproate). All 29 participants were on stable doses of their medication for at least 4 weeks prior to enrolling in the study and remained on the same dose for the duration of the trial. Participants were excluded if they had a history of any neurological or serious medical conditions, significant psychiatric co-morbidities (including substance use) or were currently pregnant. Written consent was obtained prior to undertaking any study procedures. Ethics approval was granted by Monash University and the Alfred Health Ethics Committees.

### 2.2 Design

This was a double-blind parallel randomized sham-controlled trial. Participants were randomised to receive 5 sessions of either active or sham anodal 2mA tDCS to the left dorsolateral prefrontal cortex (DLPFC) over 4 days. Fifteen participants were randomised to active stimulation, and 15 to sham stimulation. The day prior to their first tDCS session all participants underwent a comprehensive cognitive battery and clinical assessment, as well as TMS-EEG and EEG during an n-back task. Twenty-four hours after their last tDCS session participants repeated these assessments (i.e. Cognitive and clinical assessment; TMS-EEG, and EEG with n-back). The study was designed to allow for the provision of a short course of tDCS (i.e. 5tx) with all treatments and assessments occurring within a single week in order to maximise participation and minimise attrition. See Figure Two for the full study design.

**Figure 2:**
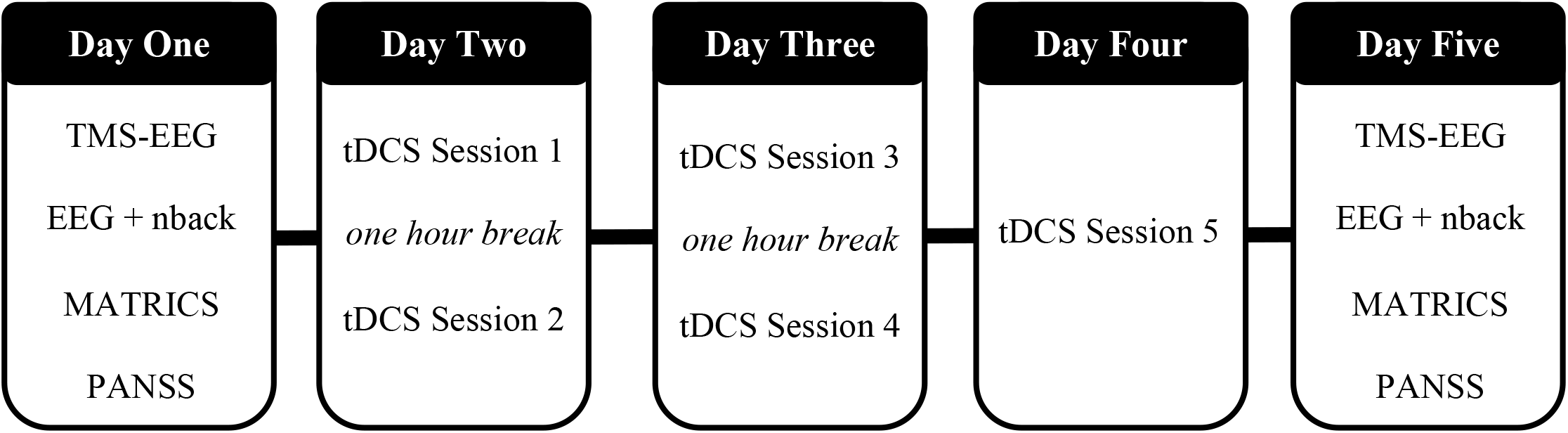
Study Protocol

### 2.3 Cognitive and Clinical Assessment

#### 2.3.1 Cognitive Assessment

Participants completed the MATRICS Consensus battery (Nuechterlein et al., 2008). The MATRICS battery is a multi-domain cognitive battery specifically designed for repeated use in people with schizophrenia. Using the MATRICS we assessed the domains of speed of processing, attention, verbal learning, working memory, visual learning, reasoning, and problem solving. Participants with schizophrenia undertook the MATRICS battery before and after the short course of tDCS.

#### 2.3.2 Clinical Measures

Participants with schizophrenia were assessed using the Positive and Negative Syndrome Scale (PANSS) (Key et al., 1987). The PANSS was undertaken before and after the short course of tDCS.

### 2.4 Neurobiological Assessments

#### 2.4.1 Transcranial Magnetic Stimulation-Electroencephalography (TMS-EEG)

TMS was delivered via a figure-of-eight MagVenture B-65 fluid-cooled coil (MagVenture A/S, Denmark) and stimulator. Resting motor threshold (RMT) was obtained from the abductor pollicis brevis (APB) muscle and was determined as the stimulus intensity required to evoke a motor evoked potential (MEP) of amplitude +50 μV in at least three out of five trials. Single pulse TMS was delivered, concurrent with EEG, at an intensity of 110% of the RMT (50 pulses, 0.25Hz plus jitter) over the left DLPFC as localised using F3 (Fitzgerald et al., 2009). EEG was recorded with a 40-scalp electrode set up using standard 10-20 positions (Quickcap, Compumedics Ltd., Australia), see Supplementary Materials for electrodes used. Electrode impedances were regularly checked and kept below 5 kΩ throughout the experiment. White noise was played through headphones to reduce auditory artefacts from the TMS pulses. A sampling rate of 10 kHz was used, and data were amplified (1000x) and online bandpass filtered (DC -2000 Hz). See Supplementary Material for details of the pre-processing and analysis of TMS-EEG data.

#### 2.4.2 EEG

EEG was recorded whilst the participants completed the 2-back working memory task at a sampling rate of 1kHz. Letters ranging from A-J appeared in the centre of the screen for 500 ms, followed by a 1500ms black screen before the next stimulus appeared. Participants responded by pressing a button if the current letter was the same as the letter from two trials beforehand. Letters were presented pseudorandomly across 130 trials (25% of which contained targets) with the task taking 5 minutes to complete. Performance on the 2-back was assessed using *d* prime and accurate reaction time. *d* prime is a discriminability index which takes into account both the ability to identify targets and to minimise false alarms, thus providing a more accurate and sensitive measure of task performance (Haatveit et al., 2010). The 2-back has been repeatedly shown to be capable of assessing working memory impairment in schizophrenia, and can generate enough correct responses to enable analysis, including using EEG data (Hoy et al., 2014, 2015, 2016).

For EEG oscillations elicited during the 2-back task, a window of 0-400ms was used for statistical analysis of theta power (4-7Hz), with the time window selected in line with previous research showing that theta increased most strongly between 0-400ms after stimulus onset for the 2-back (Hill et al., 2018). See Supplementary Material for details of the pre-processing and analysis of EEG data.

### 2.5 Transcranial Direct Current Stimulation

Current was delivered through a battery-operated current stimulator (StarStim® from Neuroelectrics) with circular electrodes (*D =* 5cm). Stimulation was applied at 0.057 mA/cm^2^ for 20mins. The anode was positioned at F3, while the cathode was placed over the contralateral supraorbital area. The position of the anode and cathode are consistent with previous research stimulating the left prefrontal cortex through tDCS, including our own previous work in schizophrenia (Hoy et al., 2014, 2015, 2016). Electric field simulation of the montage is provided in the supplementary material, including Supplementary Figure One.

Stimulation was faded in for 30 seconds before reaching the desired intensity, with a fade out period of 15 seconds at the end of stimulation. Sham tDCS was achieved by switching off stimulation after approx. 30s, which occurs within the software of the StarStim system allowing for administrator and participant blinding. The current intensity and stimulation duration all fell within established standard safety ranges (Bikson et al., 2016).

### 2.6 Statistical Analysis

Statistical analyses were performed using SPSS version 24 (IBM Corp, Armonk, NY, USA), Prism version 7 (GraphPad, San Diago, CA, USA) and the FieldTrip MATLAB toolbox. To reduce the number of comparisons, we restricted analyses to the outcome measures indicated in our baseline analysis as being potential therapeutic targets, namely cortical reactivity as indexed via TEPs (specifically, the N40 and N100), TMS-related theta oscillations and task-related theta oscillations (Hoy et al., 2021). To control for both family wise error rate and false negatives, a significance value of p < 0.01 (two-tailed) was used for all analyses, with >0.01 and <0.05 considered a trend.

In light of the small sample size, Bayesian repeated measures ANOVAs were also conducted in order to quantify the level of evidence of the null hypothesis (JASP 0.14.1). A Bayes Factor (BF10) of greater than 3 was considered to provide strong evidence for the alternative hypothesis, while a BF10 of less than 0.3 was interpreted as strong evidence for the null hypothesis (Wagenmakers et al., 2018).

#### 2.6.1 Neurobiological Data

See Supplementary Materials for details of the pre-processing and analysis of TMS-EEG and EEG data

##### 2.6.1.1 Region of Interest (ROI) Analysis

Repeated measures ANOVAs were used to investigate the effect of stimulation condition over time for N40 and N100 TEP amplitude and theta oscillatory power (from both TMS-EEG and during the 2-back EEG), extracted from a ROI corresponding to the left prefrontal cortex (electrodes F1, F3, FC1, FC3).

##### 2.6.1.2 Global Analysis

Non-parametric cluster-based permutation analyses were used to test for statistical differences in the TMS-EEG and EEG data sets (i.e. N40, N100, TMS-related theta oscillations and task-related theta oscillations). This approach allows for investigation of global differences across all electrodes, while controlling for multiple comparisons (Maris and Oostenveld, 2007). Clusters were defined as ≥ 2 neighbouring electrodes in which the t-statistic exceeded the threshold of p < 0.05 and Monte Carlo p-values (two-tailed) were subsequently calculated (1000 iterations). Analyses compared the pre- and post-tDCS timepoints within the active and the sham groups separately.

#### 2.6.2 Behavioural Data

Repeated measures ANOVAs were used to investigate the effect of stimulation conditions over time on cognitive (2-back and MATRICS) and clinical data (PANSS). The primary analyses for the behavioural data were performance on the 2-back (i.e. dprime and accurate reaction time), while exploratory analyses were conducted to investigate the impact of stimulation on the MATRICS domain scores and the PANSS subscales.

## 3. Results

### 3.1 TMS-EEG

In the active tDCS group an average of 47.63 (± 1.85) and 47.38 (± 1.41) TMS-EEG trials were included from the baseline and endpoint assessments, respectively. While in the sham stimulation group, 47.33 (± 2.29) trials were included from baseline and 46.33 (± 1.50) from endpoint.

#### 3.1.1 TMS-Evoked Potentials (TEPs)

The TEP waveforms from baseline and endpoint assessments for both the active and sham groups are shown in Figure Three. The standard four peaks obtained (N40, P60, N100, P200) are outlined (Hill et al.,2016). See Supplementary Materials for information on the Signal-to-Noise Ratio (SNR) of the data.

**Figure 3:**
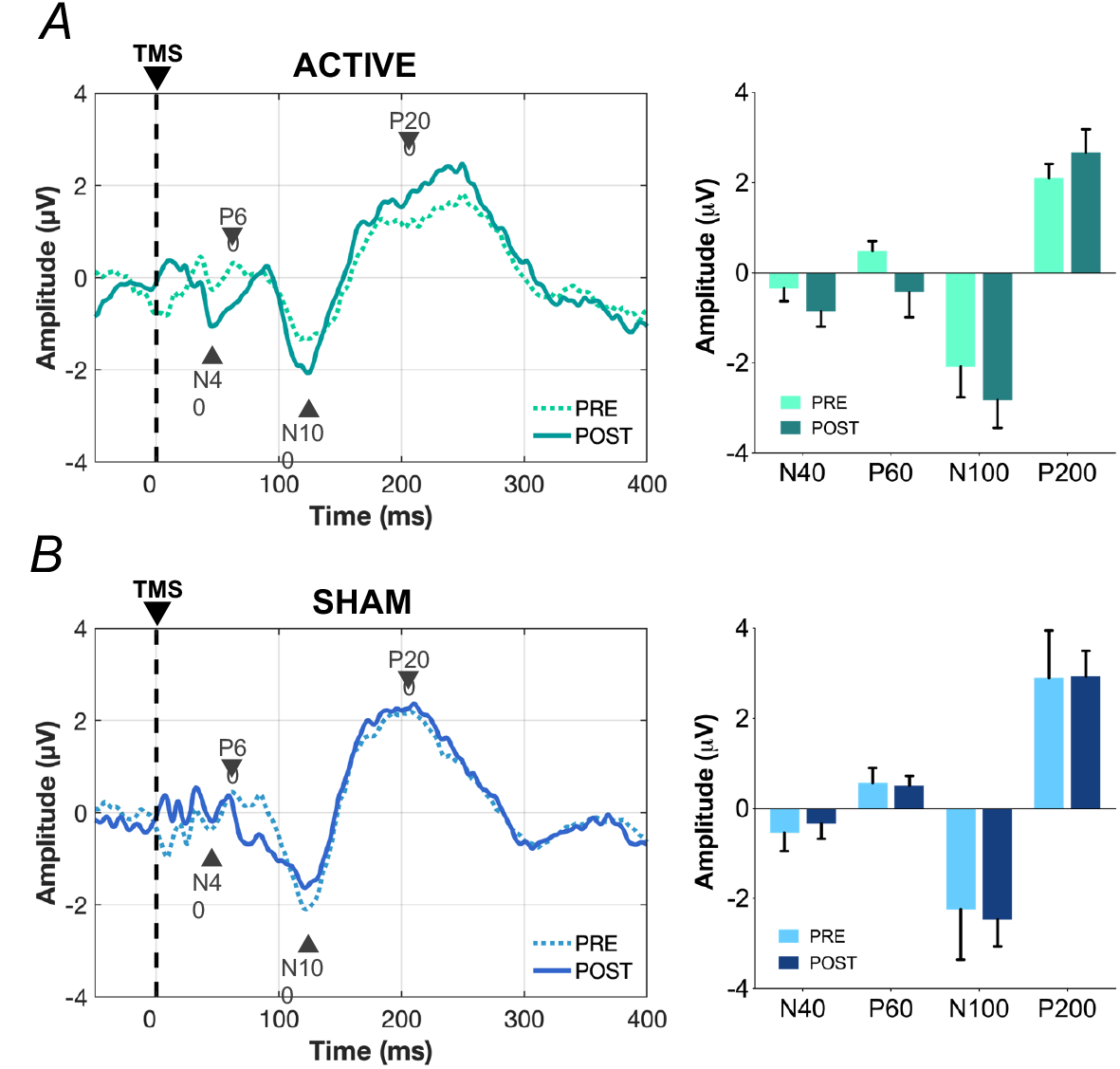
TMS-evoked potentials elicited following stimulation over the left DLPFC (TEP waveform is from the average across the left frontal ROI electrodes; F1, F3, FC3, FC1) pre- and post-active stimulation (A) and pre- and post-sham stimulation (B). The bar graphs depict the average amplitude for each TEP, error bars are SEM.

ROI comparisons were run between the baseline and endpoint timepoints for each stimulation condition for the N40 and N100 peak amplitudes. There were no significant time*group interactions for the N40 (*F*_(1,15)_ = 0.032, p=0.860; BF_10_= 0.403) or the N100 (*F*_(1,15)_ = 0.069, p=0.797; BF_10_= 0.427). See Supplementary Table 2 for means and standard deviations.

Comparison of the groups using cluster-based permutation analyses revealed that following stimulation, the active group showed a significant decrease in N40 amplitude over the left parietal-occipital region (p =0.007; with the difference most present in electrodes P5, P3, P1, PO3, PO7, O1, Oz), See Figure Four. There were no changes in the sham group.

**Figure 4:**
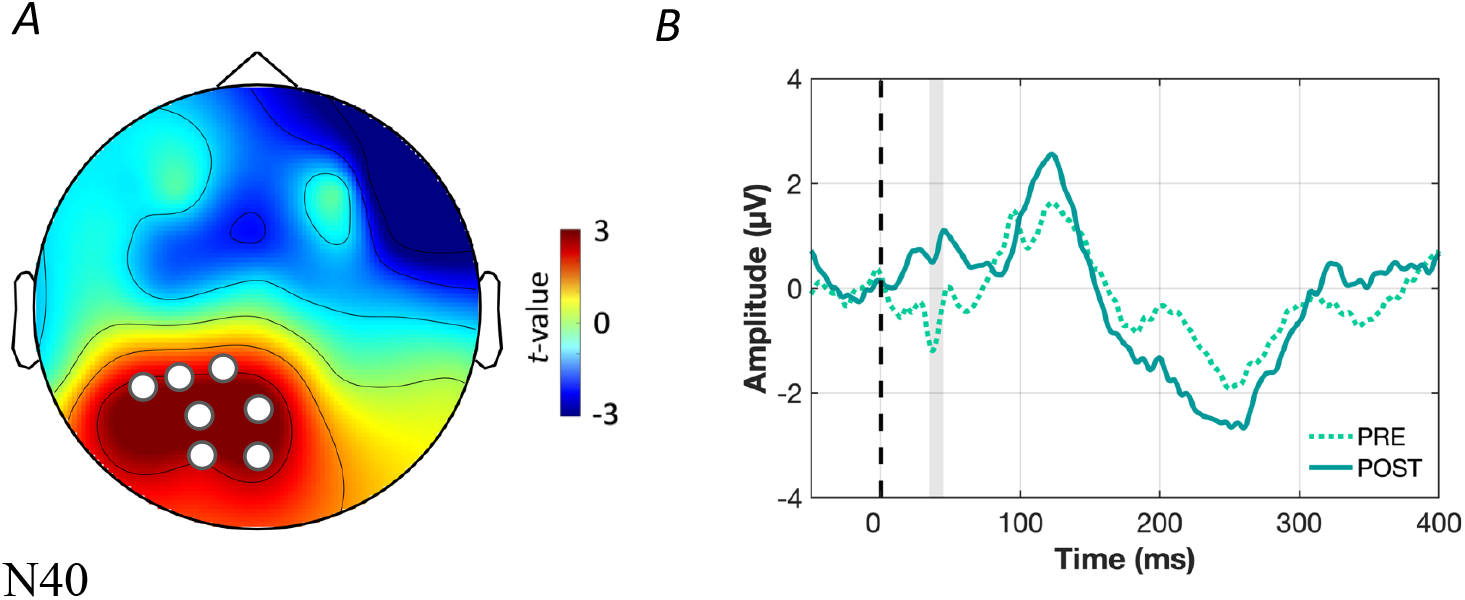
Topographical map displaying the electrodes forming significant clusters for the difference in N40 before and after active stimulation (A) and plot of the TEPs taken using the average across the electrodes forming the significant cluster (B). The dashed black line denotes the TMS pulse, while the vertical grey bar indicates the time window used for statistical analysis.

#### 3.1.2 TMS-related Oscillations

ROI analyses showed no significant TMS-related theta oscillations (*F*_(1,15)_ = 0.921, p=0.352; BF_10_= 0.831). See Supplementary Table 2 for means and standard deviations. Cluster-based permutation analyses also showed no significant changes following stimulation for either group.

### 3.2 Task-related EEG

There was no significant time*group interaction for the ROI time-frequency representations of theta oscillatory power elicited during the 2-back task (*F*_(1,21)_= 0.003, p=0.937; BF_10_= 0.385). See Supplementary Table 2 for means and standard deviations. Cluster-based permutation analyses again showed no significant changes following stimulation for either group.

With respect to behavioural performance on the 2-back, there were no significant time*group interactions for either *d* prime (*F*_(1,23)_ = 0.033, p=0.858; BF_10_= 0.375) or accurate reaction time (*F*_(1,23)_ = 0.005, p=0.943; BF_10_= 0.376), see Supplementary Table 3 for means and standard deviations.

### 3.3 Cognitive Data: MATRICS

There was a trend level time*group interaction for the reasoning and problem-solving domain, (*F*_(1,24)_ = 5.446, p = 0.028; BF_10_= 0.335). Post hoc tests showed no significant change in performance within either group over time (active: *t*_(12)_ = 1.697, p = 0.116; BF_10_= 0.618, sham: *t*_((12)_ = -1.606, p = 0.134; BF_10_= 0.656). There was a near-significant difference at baseline however, with participants in the active group performing significantly better than those in sham (*t*_(24)_ = 2.729, p = 0.012; BF_10_= 3.050). There was no difference between the groups following stimulation (*t*_(24)_= 1.369, p = 0.184; BF_10_= 0.345). There were no other significant differences between the active and sham group on any of the other MATRICS domains. See Supplementary Table 3 for means and standard deviations.

### 3.4 Clinical

With respect to clinical outcomes, there was a trend level time*group interaction for the general scale of the PANSS (*F*_(1,24)_ = 4.224, p = 0.051; BF_10_= 0.346). Post hoc analysis revealed that those in the active group showed a significant reduction in scores following stimulation (*t*_(12)_ = 4.748, p<0.001; BF_10_= 0.609), with no change in the sham group (*t*_(12)_ =0.469, p = 0.648; BF_10_= 0.634). There were no other significant differences between the active and sham group on any of the other PANSS subscales. See Supplementary Table 3 for means and standard deviations.

### 3.5 tDCS Tolerability

All participants tolerated tDCS well; there were no adverse events reported.

## 4. Discussion

In this small target engagement study we saw no changes in cortical reactivity or oscillatory activity in participants with schizophrenia following 5 sessions of tDCS. Cluster based statistics did reveal a significant reduction in left parietal occipital N40 amplitude following active stimulation only. There were no changes to cognitive performance, with the exception of a time by group interaction for the *reasoning and problem-solving* domain which was driven by differences between the groups at baseline. Finally, there was a significant reduction in the PANSS general subscale for the active stimulation group only, with no significant changes on any of the other subscales nor the PANSS total score. The PANSS general psychopathology subscale consists of a wide variety of symptoms, with the reconceptualized five factor model showing ‘general’ items are more appropriately captured across a variety of factors, including depression and cognition subscales (Rodriguez-Jimenez et al., 2013). As such, it is hard to draw any conclusions as to what a significant reduction in such a broad scale may represent.

With respect to the cortical reactivity measures, we hypothesised that tDCS would lead to increases in prefrontal TEP amplitudes, in particular in the N40 and N100. Following active tDCS we did see a significantly altered N40 in the left parietal occipital region, with the TEP plot indicating a reversal of the N40 polarity following stimulation. There were no other significant TEP changes, despite a greater mean change in amplitude towards an increased prefrontal N100 following active stimulation (mean Δ =0.74, se= 0.58) compared to sham (mean Δ= -0.23, se= 0.77). It is pertinent here to consider the posited origins of these relatively novel TMS related neurobiological outcomes. There are now several lines of evidence, including drug challenge studies, suggesting that the N100 may reflect GABA_B_ mediated cortical inhibition (Farzan et al., 2013; Premoli et al., 2013; Rogasch et al., 2013); while also likely impacted by auditory and somatosensory processing (Conde et al., 2019). While the N40 is thought to potentially be related to GABA_A_-mediated inhibition (Noda et al., 2017; Premoli et al., 2014; Voineskos et al., 2019), there is considerably less evidence for this association.

The functional significance of the N40 finding is challenging to interpret in light of the limited evidence of its cortical origin. However, from what is known, the result could suggest that active prefrontal stimulation influenced N40 activity in distal brain regions - specifically left parietal-occipital. In contrast to the largely consistent findings indicating reduced *prefrontal* cortical inhibition in schizophrenia, in other regions (including parietal-occipital regions) findings have been mixed (Egerton et al., 2017; Marsman et al., 2014; Öngür et al., 2010). The ability to differentially modulate brain activity throughout distally connected cortical regions could be advantageous for the generation of dynamic and balanced cortical activity throughout the brain, such as is required for successful cognitive functioning (Park and Friston, 2013). Irrespective of the possible functional significance, the current study does provide some limited support for the hypothesis that tDCS is able to modulate target brain activity in schizophrenia. There was, however, no modulation of either oscillatory power or cognitive performance seen in the current study. Previous research has shown that the effects of brain stimulation, and prefrontal tDCS in particular, are highly influenced by inter-individual variability (Filmer at al., 2019; Luque-Casado et al., 2019; Tremblay et al., 2014). Therefore, it is possible that those participants who show the greatest neurobiological response to stimulation are those most likely to show a behavioural response. In light of the small sample size, we were unable to explore this in the current study.

The primary limitation of the study was the small sample size, with the paucity of significant findings across the neurobiological and cognitive measures likely due to a lack of power, as was indicated the Bayesian analyses. As discussed above, there is a large and growing body of research looking at the interindividual variability of response to brain stimulation (Filmer at al., 2019; Luque-Casado et al., 2019; Tremblay et al., 2014), something that is highly likely to have impacted our pre- and post-tDCS results. However, we could not explore this further due to the current sample size. Future research should investigate engagement of evidence based therapeutic targets in large scale RCTs to confirm target engagement and relevance to symptom outcome. In addition, we only provided a short course of tDCS (5 sessions) which may have been insufficient to produce changes 24 hours following the last stimulation session when our endpoint assessments were conducted. With respect to the TMS-EEG protocol, we used 50 pulses and a higher number may have increased the data quality.

However, the number of pulses was chosen was done so to keep the data collection sessions to a manageable duration for participants, and our SNR analysis by and large showed acceptable ratios consistent with other studies of DLPFC TMS-EEG (Chung et al., 2017; Hill et al., 2019). Finally, use of the F3 to localise the left DLPFC for TMS-EEG, while shown by our previous research to provide an accurate representation (Fitzgerald et al., 2009), should be substituted with neuronavigation where possible and practical.

The current study investigated engagement of experimentally supported potential therapeutic targets for the treatment of cognitive impairment in schizophrenia using tDCS. Future interrogation of such targets in large scale RCTs will allow for the validation of both the ability of brain stimulation to modulate them and their clinical relevance. The development of testable therapeutic mechanisms for cognitive treatment development in brain stimulation is critical for the advancement of the field.

## Data Availability

The data are not publicly available due to them containing information that could compromise research participant privacy/consent.

## Acknowledgements

KEH was supported to conduct this research by the National Health and Medical Research Council (NHMRC) Fellowships (1082894 & 1135558). PBF was supported by an NHMRC Practitioner Fellowship (1078567).

---

**Supplementary Figure One:**
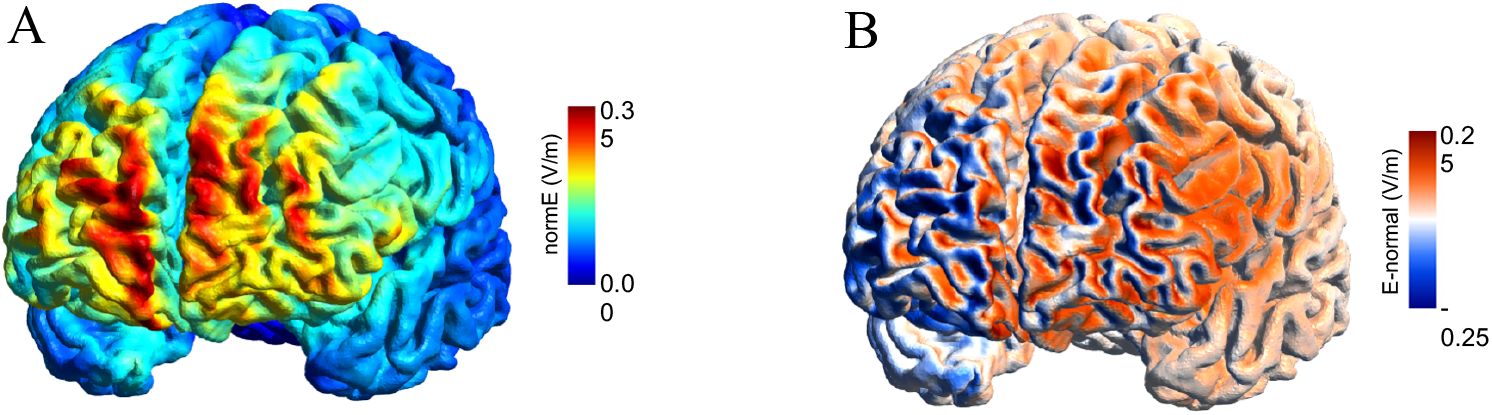

## References

Abbott, A., 2010. The drug deadlock. Nature, 468(7321), p.158.

Bikson, M., Grossman, P., Thomas, C., Zannou, A.L., Jiang, J., Adnan, T., Mourdoukoutas, A.P., Kronberg, G., Truong, D., Boggio, P. and Brunoni, A.R., 2016. Safety of transcranial direct current stimulation: evidence based update 2016. Brain stimulation, 9(5), pp.641–661.

Bora, E., Yücel, M. and Pantelis, C., 2010. Cognitive impairment in schizophrenia and affective psychoses: implications for DSM-V criteria and beyond. Schizophrenia bulletin, 36(1), pp.36–42..

Chung, S.W., Lewis, B.P., Rogasch, N.C., Saeki, T., Thomson, R.H., Hoy, K.E., Bailey, N.W. and Fitzgerald, P.B., 2017. Demonstration of short-term plasticity in the dorsolateral prefrontal cortex with theta burst stimulation: A TMS-EEG study. Clinical Neurophysiology, 128(7), pp.1117–1126.

Cirillo, G.D.P.G., Di Pino, G., Capone, F., Ranieri, F., Florio, L., Todisco, V., Tedeschi, G., Funke, K. and Di Lazzaro, V., 2017. Neurobiological after-effects of non-invasive brain stimulation. Brain stimulation, 10(1), pp.1–18.

Egerton, A., Modinos, G., Ferrera, D. and McGuire, P., 2017. Neuroimaging studies of GABA in schizophrenia: a systematic review with meta-analysis. Translational psychiatry, 7(6), pp.e1147–e1147..

Farzan, F., Barr, M.S., Hoppenbrouwers, S.S., Fitzgerald, P.B., Chen, R., Pascual-Leone, A. and Daskalakis, Z.J., 2013. The EEG correlates of the TMS-induced EMG silent period in humans. Neuroimage, 83, pp.120–134.

Filmer, H.L., Ehrhardt, S.E., Shaw, T.B., Mattingley, J.B. and Dux, P.E., 2019. The efficacy of transcranial direct current stimulation to prefrontal areas is related to underlying cortical morphology. NeuroImage, 196, pp.41–48.

Fitzgerald, P.B., Maller, J.J., Hoy, K.E., Thomson, R. and Daskalakis, Z.J., 2009. Exploring the optimal site for the localization of dorsolateral prefrontal cortex in brain stimulation experiments. Brain stimulation, 2(4), pp.234–237.

Frankle, W.G., Cho, R.Y., Prasad, K.M., Mason, N.S., Paris, J., Himes, M.L., Walker, C., Lewis, D.A. and Narendran, R., 2015. In vivo measurement of GABA transmission in healthy subjects and schizophrenia patients. American Journal of Psychiatry, 172(11), pp.1148–1159.

Gonzalez-Burgos, G. and Lewis, D.A., 2008. GABA neurons and the mechanisms of network oscillations: implications for understanding cortical dysfunction in schizophrenia. Schizophrenia bulletin, 34(5), pp.944–961.

Green, M.F., Kern, R.S. and Heaton, R.K., 2004. Longitudinal studies of cognition and functional outcome in schizophrenia: implications for MATRICS. Schizophrenia research, 72(1), pp.41–51.

Haatveit, B.C., Sundet, K., Hugdahl, K., Ueland, T., Melle, I. and Andreassen, O.A., 2010. The validity of d prime as a working memory index: results from the “Bergen n-back task”. Journal of clinical and experimental neuropsychology, 32(8), pp.871–880.

Hasan, A., Strube, W., Palm, U. and Wobrock, T., 2016. Repetitive noninvasive brain stimulation to modulate cognitive functions in schizophrenia: a systematic review of primary and secondary outcomes. Schizophrenia bulletin, 42(suppl_1), pp.S95–S109.

Herrmann, C.S., Rach, S., Neuling, T. and Strüber, D., 2013. Transcranial alternating current stimulation: a review of the underlying mechanisms and modulation of cognitive processes. Frontiers in human neuroscience, 7, p.279.

Hill, A.T., Rogasch, N.C., Fitzgerald, P.B. and Hoy, K.E., 2019. Impact of concurrent task performance on transcranial direct current stimulation (tDCS)-Induced changes in cortical physiology and working memory. Cortex, 113, pp.37–57.

Hill, A.T., Rogasch, N.C., Fitzgerald, P.B. and Hoy, K.E., 2018. Effects of single versus dual-site High-Definition transcranial direct current stimulation (HD-tDCS) on cortical reactivity and working memory performance in healthy subjects. Brain stimulation, 11(5), pp.1033–1043.

Hill, A.T., Rogasch, N.C., Fitzgerald, P.B. and Hoy, K.E., 2016. TMS-EEG: A window into the neurophysiological effects of transcranial electrical stimulation in non-motor brain regions. Neuroscience & Biobehavioral Reviews, 64, pp.175–184.

Hoy, K.E., Arnold, S.L., Emonson, M.R., Daskalakis, Z.J. and Fitzgerald, P.B., 2014. An investigation into the effects of tDCS dose on cognitive performance over time in patients with schizophrenia. Schizophrenia research, 155(1-3), pp.96–100.

Hoy, K.E., Bailey, N.W., Arnold, S.L. and Fitzgerald, P.B., 2015. The effect of transcranial Direct Current Stimulation on gamma activity and working memory in schizophrenia. Psychiatry research, 228(2), pp.191–196.

Hoy KE, Coyle, H, Gainsford K, Hill A, Bailey N, & Fitzgerald PB. Investigating neurophysiological markers of impaired cognition in schizophrenia. Schizophrenia Research 233, 34–43, 2021.

Hoy, K.E. and Fitzgerald, P.B., 2010. Brain stimulation in psychiatry and its effects on cognition. Nature Reviews Neurology, 6(5), pp.267–275.

Hoy, K.E., Whitty, D., Bailey, N. and Fitzgerald, P.B., 2016. Preliminary investigation of the effects of γ-tACS on working memory in schizophrenia. Journal of Neural Transmission, 123(10), pp.1205–1212.

Krause, B., Márquez-Ruiz, J. and Cohen Kadosh, R., 2013. The effect of transcranial direct current stimulation: a role for cortical excitation/inhibition balance?. Frontiers in human neuroscience, 7, p.602.

Insel, T.R., 2010. Rethinking schizophrenia. Nature, 468(7321), pp.187–193.

Jiang, Y., Guo, Z., Xing, G., He, L., Peng, H., Du, F., McClure, M.A. and Mu, Q., 2019. Effects of high-frequency transcranial magnetic stimulation for cognitive deficit in schizophrenia: a meta-analysis. Frontiers in psychiatry, 10, p.135.

Kay, S.R., Fiszbein, A. and Opler, L.A., 1987. The positive and negative syndrome scale (PANSS) for schizophrenia. Schizophrenia bulletin, 13(2), pp.261–276.

Lisman, J., 2012. Excitation, inhibition, local oscillations, or large-scale loops: what causes the symptoms of schizophrenia?. Current opinion in neurobiology, 22(3), pp.537–544.

Luque-Casado, A., Fogelson, N., Iglesias-Soler, E. and Fernandez-del-Olmo, M., 2019. Exploring the effects of Transcranial Direct Current Stimulation over the prefrontal cortex on working memory: A cluster analysis approach. Behavioural brain research, 375, p.112144.

Maris, E. and Oostenveld, R., 2007. Nonparametric statistical testing of EEG-and MEG-data. Journal of neuroscience methods, 164(1), pp.177–190.

Marsman, A., Mandl, R.C., Klomp, D.W., Bohlken, M.M., Boer, V.O., Andreychenko, A., Cahn, W., Kahn, R.S., Luijten, P.R. and Pol, H.E.H., 2014. GABA and glutamate in schizophrenia: A 7 T 1H-MRS study. NeuroImage: Clinical, 6, pp.398–407.

Mervis, J.E., Capizzi, R.J., Boroda, E. and MacDonald III, A.W., 2017. Transcranial direct current stimulation over the dorsolateral prefrontal cortex in schizophrenia: a quantitative review of cognitive outcomes. Frontiers in Human Neuroscience, 11, p.44..

Narita, Z., Stickley, A., DeVylder, J., Yokoi, Y., Inagawa, T., Yamada, Y., Maruo, K., Koyanagi, A., Oh, H., Sawa, A. and Sumiyoshi, T., 2020. Effect of multi-session prefrontal transcranial direct current stimulation on cognition in schizophrenia: A systematic review and meta-analysis. Schizophrenia research, 216, pp.367–373.

Noda, Y., Zomorrodi, R., Cash, R.F., Barr, M.S., Farzan, F., Rajji, T.K., Chen, R., Daskalakis, Z.J. and Blumberger, D.M., 2017. Characterization of the influence of age on GABAA and glutamatergic mediated functions in the dorsolateral prefrontal cortex using paired-pulse TMS-EEG. Aging (Albany NY), 9(2), p.556..

Nuechterlein, K.H., Green, M.F., Kern, R.S., Baade, L.E., Barch, D.M., Cohen, J.D., Essock, S., Fenton, W.S., Frese III, Ph D, F.J., Gold, J.M. and Goldberg, T., 2008. The MATRICS Consensus Cognitive Battery, part 1: test selection, reliability, and validity. American Journal of Psychiatry, 165(2), pp.203–213.

Öngür, D., Prescot, A.P., McCarthy, J., Cohen, B.M. and Renshaw, P.F., 2010. Elevated gamma-aminobutyric acid levels in chronic schizophrenia. Biological psychiatry, 68(7), pp.667–670.

Park, H.J. and Friston, K., 2013. Structural and functional brain networks: from connections to cognition. Science, 342(6158).

Premoli, I., Castellanos, N., Rivolta, D., Belardinelli, P., Bajo, R., Zipser, C., Espenhahn, S., Heidegger, T., Müller-Dahlhaus, F. and Ziemann, U., 2014. TMS-EEG signatures of GABAergic neurotransmission in the human cortex. Journal of Neuroscience, 34(16), pp.5603–5612.

Rodriguez-Jimenez, R., Bagney, A., Mezquita, L., Martinez-Gras, I., Sanchez-Morla, E.M., Mesa, N., Ibañez, M.I., Diez-Martin, J., Jimenez-Arriero, M.A., Lobo, A. and Santos, J.L., 2013. Cognition and the five-factor model of the positive and negative syndrome scale in schizophrenia. Schizophrenia Research, 143(1), pp.77–83.

Rogasch, N.C., Daskalakis, Z.J. and Fitzgerald, P.B., 2013. Mechanisms underlying long-interval cortical inhibition in the human motor cortex: a TMS-EEG study. Journal of neurophysiology, 109(1), pp.89–98.

Schmiedt, C., Brand, A., Hildebrandt, H. and Basar-Eroglu, C., 2005. Event-related theta oscillations during working memory tasks in patients with schizophrenia and healthy controls. Cognitive Brain Research, 25(3), pp.936–947.

Shamsi, S., Lau, A., Lencz, T., Burdick, K.E., DeRosse, P., Brenner, R., Lindenmayer, J.P. and Malhotra, A.K., 2011. Cognitive and symptomatic predictors of functional disability in schizophrenia. Schizophrenia research, 126(1-3), pp.257–264.

Sheehan, D.V., Lecrubier, Y., Sheehan, K.H., Amorim, P., Janavs, J., Weiller, E., Hergueta, T., Baker, R. and Dunbar, G.C., 1998. The Mini-International Neuropsychiatric Interview (MINI): the development and validation of a structured diagnostic psychiatric interview for DSM-IV and ICD-10. The Journal of clinical psychiatry.

Sohal, V.S. and Rubenstein, J.L., 2019. Excitation-inhibition balance as a framework for investigating mechanisms in neuropsychiatric disorders. Molecular psychiatry, 24(9), pp.1248–1257.

Tabachnick, B.G., Fidell, L.S. and Ullman, J.B., 2007. Using multivariate statistics (Vol. 5, pp. 481–498). Boston, MA: Pearson.

Tremblay, S., Lepage, J.F., Latulipe-Loiselle, A., Fregni, F., Pascual-Leone, A. and Théoret, H., 2014. The uncertain outcome of prefrontal tDCS. Brain stimulation, 7(6), pp.773–783..

Veniero, D., Vossen, A., Gross, J. and Thut, G., 2015. Lasting EEG/MEG aftereffects of rhythmic transcranial brain stimulation: level of control over oscillatory network activity. Frontiers in cellular neuroscience, 9, pp.477.

Voineskos, D., Blumberger, D.M., Zomorrodi, R., Rogasch, N.C., Farzan, F., Foussias, G., Rajji, T.K. and Daskalakis, Z.J., 2019. Altered transcranial magnetic stimulation–electroencephalographic markers of inhibition and excitation in the dorsolateral prefrontal cortex in major depressive disorder. Biological psychiatry, 85(6), pp.477–486.-486.

Wagenmakers EJ, Love J, Marsman M, Jamil T, Ly A, Verhagen J, Selker R, Gronau QF, Dropmann D, Boutin B, Meerhoff F., 2018. Bayesian inference for psychology. Part II: Example applications with JASP. Psychonomic bulletin & review, 25(1), pp 58–76.

Wykes, T., Huddy, V., Cellard, C., McGurk, S.R. and Czobor, P., 2011. A meta-analysis of cognitive remediation for schizophrenia: methodology and effect sizes. American Journal of Psychiatry, 168(5), pp.472–485.

