## Supplementary Materials for "Investigating neurophysiological effects of a short course of tDCS for cognition in schizophrenia: a target engagement study"

**Methods**

***Neurobiological Assessments***

*Electrode Used*: AF3, AF4, F7, F5, F3, F1, FZ, F2, F4, F6, F8, FC5, FC3, FC1, FCZ, FC2, FC4, FC6, C5, C3, C1, CZ, C2, C4, C6, P7, P5, P3, P1, PZ, P2, P4, P6, P8, PO3, POZ, PO4, O1, OZ, O2, M1, M2

*TMS-EEG data pre-processing*

TMS pulses were identified within the EEG trace using the '*tesa_findpulsepeak'* function from the TESA software platform (TMS-EEG Signal Analyser [Rogasch et al., 2017]). The data were then segmented into two-second epochs (-1000 to 1000 ms) around each TMS pulse and baseline corrected (-500 to -110 ms). A -5 to 10 ms time-window containing the TMS pulse and associated large amplitude artefact was then removed and replaced using linear interpolation using the *'pop_tesa_interpdata'* function in the TESA software (i.e., linear function fitted on data point before and after the missing data). The data were then down-sampled from 10 KHz to 1 KHz. Any excessively noisy channels/trials were then removed based on visual inspection, following which an initial round of independent component analysis (ICA; FastICA algorithm [Hyvärinen and Oja, 2000]) was performed to remove components containing any residual high amplitude TMS-evoked EMG artefact. Data were then band-pass (1-100 Hz) and notch filtered (48-52 Hz, to reduce line noise) using 4$^{th}$ order zero-phase Butterworth filters. Data were again visually inspected, and any remaining noisy channels/trials removed. ICA was then performed for a second time to eliminate any remaining smaller components representing blink, eye movement, persistent muscle activity and electrode noise. Missing channels were then replaced using spherical interpolation and data were re-referenced to the common average.

*EEG data pre-processing*

EEG data were band-pass (0.5 - 100 Hz) and band-stop (48-52 Hz) filtered using a 4$^{th}$-order zero phase Butterworth filter, before being segmented (-1500 – 1999 ms) around stimulus onset for correctly encoded trials (encoding trials where the subsequent probe trial was responded to correctly). Any epochs containing a keypad press within -500 to 1000 ms of the stimulus onset were also removed to prevent any confounds of motor-related EEG changes. The data were then baseline corrected (-300 to 0ms) and visually inspected, with any excessively noisy channels/trials removed. ICA (FastICA algorithm [Hyvärinen and Oja, 2000]) was then performed to remove any additional artefacts, following which the EEG trace was again reviewed and any remaining bad channels/trials removed. Any missing channels were replaced using spherical interpolation and data were referenced to the common average.

***Data Analysis***

*TMS-Evoked Potentials*

Single-pulse TMS over the DLPFC produces a series of positive and negative deflections with individual latencies at approximately 40ms (N40), 60ms (P60), 100ms (N100) and 200ms (P200), respectively (Farzan et al., 2013; Rogasch et al., 2015). TEPs were first calculated by averaging across the single trial data, within each participant and condition using the '*tesa_tepextract'* function. The '*tesa_peakanalysis'* function was then used to identify each of the TEP peaks of interest (N40, P60, N100, P200) (Rogasch et al., 2017). Positive peaks were defined as a data point that is greater than 5 points either side of the peak, and negative peaks were defined as a data point that is more negative than 5 points either side of the peak (Hill et al., 2017). The time windows chosen for detection of the negative N40 and N100 peaks were 25-55 ms and 90-150 ms, respectively; while the positive P60 and P200 peaks were detected between 45-75 ms and 160-240 ms, respectively, similar to previous investigations (Chung et al., 2017; Hill et al., 2017). In instances where peaks were not detected within the allotted time windows, amplitudes were extracted from the centre of the pre-defined latency windows (Rogasch et al., 2017). A 10 ms window around these peak latencies, corresponding to the maximum amplitude of each peak (+/- 5ms), was then used for any statistical analysis of either the ROI (t-tests) or for global analyses across all scalp electrodes using cluster-based permutation statistics. In light of our hypothesised therapeutic targets, and to reduce comparisons, our analyses were restricted to the N40 and N100 (Hoy et al, in submission).

*Time-Frequency Analysis*

Time-frequency representations of oscillatory power for the TMS-EEG and EEG data were conducted using complex Morlet wavelet decompositions. A linearly spaced number of cycles from 3 (at 4Hz) to 8 (at 60Hz) (steps of 1Hz) was used to adjust for temporal and frequency precision as a function of the frequency of the wavelet (Roach et al., 2008). Baseline normalisations were applied (-500 to -200ms) using the decibel (dB) conversion, which helps remove scale differences between frequencies/individuals and is robust to $1/f$ power scaling (Roach et al., 2008). Power was analysed for the theta (4-7Hz) frequency band. Prior to statistical analyses, power values were averaged across the time window of interest, and across the theta frequency band.

For TMS-related oscillations, time windows were 10-300ms for theta frequencies. For EEG oscillations elicited during the 2-back task, a window of 0-400ms was used for theta, encapsulating the large event-related synchronisation response that occurs early in the 2-back epochs. Again, our analyses were restricted to the theta frequency band in light of our hypothesised therapeutic targets, and to reduce the number of comparisons (Hoy et al, 2021).

***Transcranial Direct Current Stimulation***

*Electric Field Modelling of the tDCS montage*

For electric field simulation of the tDCS montage, both the overall E-field strength and the normal component of the E-field (i.e., orthogonal to the cortex), were computed using the SimNIBS software platform (version 3.0.5 [Windhoff et al., 2013]). As individual subject MRI scans were not available, models of cortical current flow were created using the 'Ernie' MRI-derived template head model included with the SimNIBS software. The montage was simulated using 2 mm thick circular electrodes with a diameter of 5 cm, encased in saline soaked sponges. The head mesh used in the model was partitioned into layers which were assigned the following electrical conductivities: Scalp=0.465 S/m, skull=0.010 S/m, CSF=1.654 S/m, grey matter=0.275 S/m, white matter=0.126 S/m. Post-processing and visualization of electrical fields was performed using Gmsh (Geuzaine and Remacle, 2009).  The montage shows bi-hemispheric spread, with the normal component of the E-field showing the left hemisphere receives predominantly anodal currents, see Supplementary Figure 2.

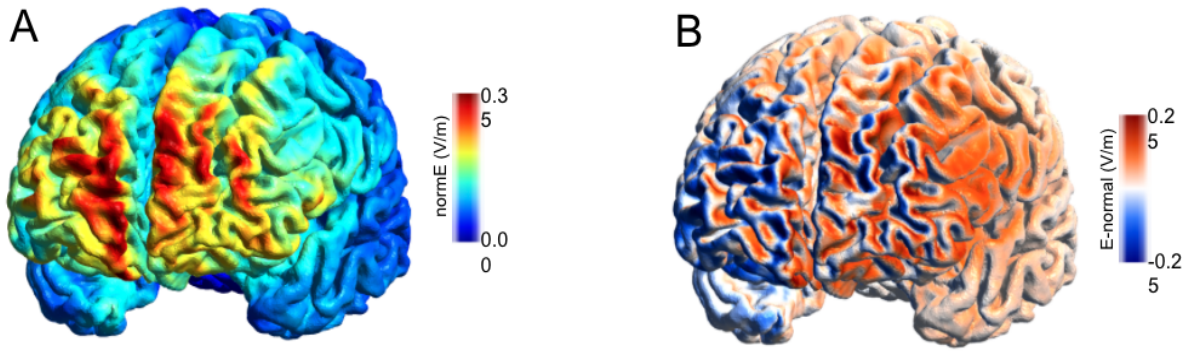


**Supplementary Figure One**: Simulation of the tDCS electric field. A) Electric field strength. B) Normal component of the electric field. Positive (red) values indicate current flowing into the cortex, while negative values (blue) indicate current flowing out of the cortex.

**Results**

*TMS-EEG*

Signal-to-noise (SNR) was performed for each TEP peak (ROI electrodes) separately across the timepoints. SNR was estimated by the standard deviation of the signal amplitude in the pre-stimulus interval (-500 to 100ms). This provides an estimate (in SD units) of the peak signal relative to the background EEG. Values greater than 3 SDs (99.7% of the baseline distribution) are generally considered to have acceptable SNR (Chung et al., 2017; Hill et al., 2017). See Supplementary Table 1 for SNR values.

Supplementary Table 1. SNR values for each TEP at the two timepoints.

| | Baseline | | End | |
|---|---|---|---|---|
| | **Mean** | **SD** | **Mean** | **SD** |
| N40 | -3.37 | 7.64 | -2.30 | 3.88 |
| P60 | 3.98 | 6.11 | 0.28 | 4.86 |
| N100 | -16.24 | 20.34 | -10.38 | 6.82 |
| P200 | 18.99 | 17.54 | 11.07 | 6.09 |

*TMS-EEG and EEG Data*

Supplementary Table 2. Means and standard deviations for TMS-EEG and EEG ROI data pre and post stimulation as a function of stimulation group

| | **Active** | | **Sham** | |
|---|---|---|---|---|
| | **Baseline** | **End** | **Baseline** | **End** |
| **TEPs** | ***Mean ± sd*** | ***Mean ± sd*** | ***Mean ± sd*** | ***Mean ± sd*** |
| N40 Amplitude | -0.349 ± 0.797 | -0.862 ± 0.942 | 0.038 ± 1.027 | -0.338 ± 1.011 |
| N100 Amplitude | -2.081 ± 1.928 | -2.854 ± 1.751 | -1.983 ± 3.563 | -2.462 ± 1.798 |
| **TMS-related Oscillations** | | | | |
| TMS-related theta oscillations | 0.893± 1.227 | 1.177 ± 1.567 | 1.773 ± 2.821 | 1.536 ± 2.057 |
| **Task-related Oscillations** | | | | |
| Task-related theta oscillations | 0.694 ± 1.328 | 0.577 ± 1.738 | 1.173 ± 1.056 | 1.087 ± 0.995 |

*Clinical and Cognitive Data*

Supplementary Table 3. Means, standard deviations and p-statistics for performance on cognitive tasks pre and post stimulation as a function of stimulation group.

| | **Active** | | **Sham** | | |
|---|---|---|---|---|---|
| | **Baseline** | **End** | **Baseline** | **End** | |
| **MATRICS Cognitive domain** | ***Mean ± sd*** | ***Mean ± sd*** | ***Mean ± sd*** | ***Mean ± sd*** | ***interaction*** |
| Speed of information processing | 36.92 ± 10.46 | 40.62 ± 10.48 | 39.77 ± 10.84 | 45.69 ± 11.79 | p = 0.254 |
| Attention/Vigilance | 36.08 ± 10.20 | 40.08 ± 11.84 | 42.33 ± 12.07 | 43.33 ± 15.28 | p = 0.310 |
| Working Memory | 40.31 ± 10.62 | 43.08 ± 11.11 | 43.15 ± 13.00 | 44.38 ± 15.47 | p =0.550 |
| Verbal Learning | 38.85 ± 12.02 | 37.15 ± 9.62 | 42.46 ± 7.26 | 38.23 ± 7.190 | p = 0.422 |
| Visual Learning | 38.92 ± 15.04 | 41.77 ± 11.41 | 39.08 ± 14.45 | 42.69 ± 11.41 | p = 0.841 |
| Reasoning and Problem solving | 49.38 ± 9.13 | 46.85 ± 9.06 | 40.46 ± 7.45 | 42.54 ± 9.06 | p =0.028* |
| **2back** | | | | | |
| Accurate reaction time (ms) | 709.08 ± 195.20 | 681.29 ± 184.98 | 730.50 ± 132.68 | 698.92 ± 183.160 | p =0.943 |
| *d* prime | 2.03 ± 0.95 | 2.55 ± 1.87 | 2.45 ± 0.83 | 3.03 ± 0.99 | p =0.858 |
| **Clinical Scales** | | | | | |
| PANSS positive | 18.85 ± 6.98 | 17.54 ± 7.37 | 19.08 ± 4.010 | 17.46 ± 3.64 | p =0.671 |

| | | | | | |
|---|---|---|---|---|---|
| PANSS negative | 17.23 ± 3.60 | 17.08 ± 4.76 | 15.46 ± 3.59 | 15.62 ± 3.50 | p =0.761 |
| PANSS general | 39.38± 7.74 | 35.62 ± 8.43 | 36.92 ± 9.55 | 36.31 ± 8.10 | p =0.051^ |
| PANSS total | 74.56 ± 15.14 | 70.23 ± 18.76 | 71.46 ± 14.89 | 69.38 ± 13.25 | p =0.183 |

**Supplementary References**


Chung, S.W., Lewis, B.P., Rogasch, N.C., Saeki, T., Thomson, R.H., Hoy, K.E., Bailey, N.W. and Fitzgerald, P.B., 2017. Demonstration of short-term plasticity in the dorsolateral prefrontal cortex with theta burst stimulation: A TMS-EEG study. Clinical Neurophysiology, 128(7), pp.1117-1126.

Farzan, F., Barr, M.S., Hoppenbrouwers, S.S., Fitzgerald, P.B., Chen, R., Pascual-Leone, A. and Daskalakis, Z.J., 2013. The EEG correlates of the TMS-induced EMG silent period in humans. Neuroimage, 83, pp.120-134.

Geuzaine, C. and Remacle, J.F., 2009. Gmsh: A 3-D finite element mesh generator with built-in pre-and post-processing facilities. International journal for numerical methods in engineering, 79(11), pp.1309-1331.

Hill, A.T., Rogasch, N.C., Fitzgerald, P.B. and Hoy, K.E., 2017. Effects of prefrontal bipolar and high-definition transcranial direct current stimulation on cortical reactivity and working memory in healthy adults. Neuroimage, 152, pp.142-157.

Hyvärinen, A. and Oja, E., 2000. Independent component analysis: algorithms and applications. Neural networks, 13(4-5), pp.411-430.

Roach, B.J. and Mathalon, D.H., 2008. Event-related EEG time-frequency analysis: an overview of measures and an analysis of early gamma band phase locking in schizophrenia. Schizophrenia bulletin, 34(5), pp.907-926..

Rogasch, N.C., Daskalakis, Z.J. and Fitzgerald, P.B., 2015. Cortical inhibition of distinct mechanisms in the dorsolateral prefrontal cortex is related to working memory performance: a TMS–EEG study. Cortex, 64, pp.68-77.

Rogasch, N.C., Sullivan, C., Thomson, R.H., Rose, N.S., Bailey, N.W., Fitzgerald, P.B., Farzan, F. and Hernandez-Pavon, J.C., 2017. Analysing concurrent transcranial magnetic stimulation and electroencephalographic data: A review and introduction to the open-source TESA software. Neuroimage, 147, pp.934-951.

Windhoff, M., Opitz, A. and Thielscher, A., 2013. Electric field calculations in brain stimulation based on finite elements: an optimized processing pipeline for the generation and usage of accurate individual head models. Human brain mapping, 34(4), pp.923-935.

**Supplementary Figure Legends**

**Supplementary Figure One:** Simulation of the tDCS electric field. A) Electric field strength. B) Normal component of the electric field. Positive values indicate current flowing into the cortex, while negative values indicate current flowing out of the cortex.
